# How is physical healthcare experienced by staff, service users, and carers in adult community mental health services in a South London Mental Health Trust? A Service Evaluation

**DOI:** 10.1101/2023.01.05.23284227

**Authors:** Gracie Tredget, Julie Williams, Ray McGrath, Euan Sadler, Fiona Gaughran, Karen Ang, Natalia Stepan, Sean Cross, John Tweed, Lia Orlando, Nick Sevdalis, Integrating our Mental and Physical Healthcare Systems (IMPHS) Study Team

## Abstract

**Background:** Adults with a serious mental illness (SMI) are at greater risk of physical health morbidity and premature death than the general population, largely as a result of preventable physical health issues. Staff working in mental health services have a role to play in addressing these inequalities, but little is known about how they perceive their role and how this impacts on their practice. Understanding this better would enable services to improve their approach and support better health outcomes for SMI patients. A service evaluation was undertaken to investigate how physical healthcare is approached within adult community mental health teams (CMHTs) at a South London (UK) Mental Health Trust.

**Methods:** This was a prospective, cross-sectional evaluation design. Interviews and focus groups were conducted with clinical staff, service users and carers, to understand their experiences and to identify key barriers and facilitators to supporting physical healthcare support for adults with SMI. Thematic analysis was conducted to identify key themes which were classified into five main categories.

**Results:** 50 participants took part in the study, 38 were clinical staff, eight were service users and four were carers. We found staff widely recognised the importance of supporting physical healthcare. However, there was variability in how staff approached physical healthcare in routine practice, and differences in how physical healthcare is experienced by service users and carers. Staff were keen to engage in changes to the way physical healthcare is delivered in CMHTs. However, they sought clearer guidance on their roles and responsibilities, and wanted to better understand the rationale for changes in community mental health practice, such as increased screening for physical healthcare. Service users and carers felt equally that the role of CMHTs in physical healthcare was unclear, which limited their ability to access it and understand the benefit for their overall care. Staff articulated gaps in leadership and training that impacted on their ability to implement the overall vision for physical healthcare within the Trust.

**Conclusion:** Mental health staff recognise the role they play in supporting the physical health of adults living with SMI. This evaluation provides insight into common barriers and facilitators faced by staff, service users and carers when providing or accessing physical healthcare within adult CMHTs. These findings indicate a more comprehensive and better articulated approach to physical healthcare in mental health Trusts is needed to ensure service users and their carers understand what support is available and how to access it and to equip staff to provide and sustain that care in routine practice.

## 1 Introduction

Adults with a serious mental illness (SMI) such as schizophrenia, schizoaffective disorder, bipolar disorder, and depressive psychosis are at greater risk of physical health morbidity and premature death than the general population (1). Several small-scale UK studies have considered the challenges in providing physical healthcare for adults living with SMI. One study identified multiple barriers including mental health professionals seeing physical healthcare planning as secondary to mental health care planning (2) and another identified that processes set up for screening and managing physical healthcare for adults with SMI are often limited due to staff lacking appropriate skills and knowledge to do this (3). Both studies suggested that effective integrated care is facilitated when service users are involved in developing services and when general practitioners are aware of any secondary care planning involving the physical health of their patients.

Other research has explored the experience of mental healthcare staff who provide physical healthcare and the impact this has on the provision of care for service users. A survey of mental healthcare professionals undertaken by Papachristou et al (2019) (4) suggested that clinical staff were unclear about which service users had comorbid health conditions and frequently lack training or resources to facilitate dual care (4). Butler (2020) (5) investigated the attitudes of healthcare professionals to the provision of physical healthcare in CMHTs and suggested that how staff experience the changes and how these impact on service users’ needs to be better understood by policymakers. They also found that clinicians with specific physical health training were more likely to advocate for physical health support for their service users.

Several papers have explored the perspectives of service users (e.g., 6,7), who largely feel their physical health is not prioritised when accessing mental health services and want to see greater physical health knowledge amongst mental? healthcare professionals. Hughes (2009) (7) explored the experiences of service users with sexual health concerns and found they valued being able to talk openly about concerns relating to all aspects of their health with mental healthcare professionals and for those concerns to be addressed in a coordinated way.

Finally, a study by Onwumere et al (2018) (8) explored the experiences of carers, revealing frustrations with the lack of coordinated care for adults with SMI when moving between parts of the care system, and the systemic burden it creates for carers. They found that the identification and management of physical health problems, gaps in services for comorbid health problems and the impact on carers when supporting loved ones outside of statutory care were key concerns.

### 1.1 Rationale and aims for a service evaluation

This service evaluation was undertaken in the South London and Maudsley NHS Foundation Trust (referred to as ‘the Trust’ throughout). The National Institute for Health and Care Excellence (NICE, 2014) (9) issued guidance in 2014 for the provision of annual physical health checks for adults with SMI with care shared between primary and secondary services. The Trust responded to this guidance by developing a strategy that outlined the responsibilities of the organisation and its staff when providing physical healthcare for adults with SMI. The Trust were keen to understand how physical healthcare is currently approached by clinical staff working within adult CMHTs, and how it is experienced by service users and carers using those services.

The scope of this service evaluation was discussed and agreed between the researchers and the Trust. We aimed to gather a wide range of staff perspectives to obtain a general view and not to draw comparisons between professional groups, teams or services within the organisation.

We aimed to explore which barriers and facilitators contribute to five main areas of interest as identified by the Trust:

i. The approach and practice of staff towards physical healthcare
ii. The use of physical health systems and tools
iii. The physical health knowledge and skills used by staff
iv. The perceptions and attitudes of staff towards physical healthcare within the Trust
v. The experiences and outcomes of service users and carers

The service evaluation was designed to enable the Trust to use the insights gained to inform local decision-making and improve future routine practice regarding physical healthcare (10).

## 2 Methodology

### 2.1 Design

This was a prospective, cross-sectional service evaluation designed by the research team in collaboration with the Trust, and experts with lived and clinical experience of SMI. Qualitative data collection methods were used to gain in-depth, detailed perspectives on the evaluation questions from the study participants. We conducted interviews and focus groups with staff working in or responsible for CMHTs, as well as focus groups with service users and their carers.

### 2.2 Ethics

Approval was obtained from Clinical and Information Governance professional leads at the Trust (on 14th March 2022). All participants involved in the study were briefed prior to taking part and given written information about how their data would be used within the evaluation process and as part of any final publications. Participants provided written informed consent to participate in the study. Participation was voluntary and withdrawal was possible at any stage.

### 2.3 Setting

This service evaluation was conducted in Adult Community Mental Health services at the South London and Maudsley NHS Foundation Trust, the largest mental health Trust in the UK, serving a local population of 1.3 million people in south east London. The Trust supports approximately 40,000 service users within community services across four boroughs: Southwark, Lambeth, Lewisham, and Croydon (11).

### 2.4 Participants

#### 2.4.1 Inclusion criteria

*Staff:* were over 18 years old, currently working within adult CMHTs with experience of supporting adults with SMI with associated physical health problems.

*Service users and carers*: had to have used adult CMHT services (as a service user or carer) at the Trust within the past 12 months and be aged over 18 years old.

#### 2.4.2 Sampling and recruitment

A purposive sampling technique was used to ensure participants were from a wide range of clinical roles, from different sociodemographic groups and to ensure service user and carer representation.

##### 2.4.2.1. Clinical staff

Key clinical roles based within adult CMHTs were identified with clinical service leads. These roles included community matrons, general managers, clinical service leads, team leaders, doctors at different grades (i.e., attending level physicians and residents in the USA), nurses, social workers, and occupational therapists. Staff demographic data from the Trust were used to ensure the final sample was demographically representative of the Trust’s workforce, as set out in the 2021-2022 Workforce Equality and Diversity Report (12).

##### 2.4.2.2. Service users and carers

Service users and carers were recruited via existing patient and public involvement within the Trust. An online participation advert was also used. Service users and carers were selected using the INCLUDE framework (13) to ensure sufficient diversity and inclusion across participants. Demographic data from across all four boroughs were used to inform participant selection to ensure representativeness, using the 2021/2022 Trust-wide Equality Information Report (14). Service users and carers were paid for their involvement in accordance with national guidelines.

### 2.5. Procedure for interviews and focus groups

Participants interested in taking part were contacted by a researcher (GT) who explained the evaluation to them. All participants taking part provided written informed consent and were asked to complete a demographic questionnaire prior to the interview or focus group.

#### 2.5.1. Interview and focus group design

The evaluation team developed a schedule of questions to use in interviews and focus groups (see additional material 1). Based on the five main areas of interest (see section 1.1) the researchers co-developed questions with an independent panel of clinical staff, service users, and carers. Questions were reviewed by the evaluation team made up of clinicians and academics (including authors: GT, RM, JW, NSt, NS, FG, ES and KA). A semi-structured interview format was used, with additional prompts used to support wider enquiry during questioning. Before data collection, two pilot interviews and two pilot focus groups were undertaken to test questions and practice facilitation. All recordings were saved with a participant ID number transcribed by an independent transcriber. Transcripts were saved securely in a password protected file on a SLaM electronic drive and the original recordings were deleted.

#### 2.5.2. Interviews

General managers, clinical service leads, team leaders, mental health nurses, occupational therapists, social workers, advanced practitioners, and physical health leads were invited to participate in online interviews using Microsoft Teams. All interviews lasted for one hour.

#### 2.5.3. Focus groups

Focus groups were conducted online using Microsoft Teams with doctors, care coordinators, service users, and carers. All focus groups lasted for one hour for clinical staff, and two hours for service users and carers. Sessions were led by two facilitators (one researcher, GT, and one expert by experience) and had a maximum of six participants per group.

## 3 Stakeholder involvement

Throughout the evaluation, progress was reviewed in the existing Trust and project forums including fortnightly meetings with Physical Health Leads from the Trust and monthly evaluation team meetings.

An independent group of clinical advisors was recruited to ensure the experiences of clinical professionals and people using services were included throughout. The group was made up of clinical advisors working within Trust CMHT services (two doctors and four care coordinators) and three experts with lived experience (two service users and one carer).

Applications for expert roles were advertised and all advisors were selected following an interview. Members of the above groups were involved throughout the study to routinely share progress and gather feedback at key stages.

## 4 Analysis

The analysis was undertaken by a team with one main coder (GT), a main reviewer (JW) and two additional reviewers (ES and RM).

### 4.1 Data extraction and coding

All transcripts were uploaded to NVivo (version 12) (QSR International, 2020). Initially GT, JW and RM read three early transcripts and extracted 5 preliminary themes and related subthemes from these (see Figure 1). Once these preliminary codes were agreed, GT continued to extract and categorise codes from the remaining transcripts. JW systematically read through the detailed coding once completed to check coding quality and accuracy. ES and RM reviewed the coding structure at regular intervals: when 10%, 25%, 50%, 75% and finally 100% of transcripts had been coded. Each review interval provided an opportunity to review the original aims of the study, to review the code structure and to observe patterns from the data. The research team were then able to synthesise the data into five main themes and related subthemes.

**Figure 1.**
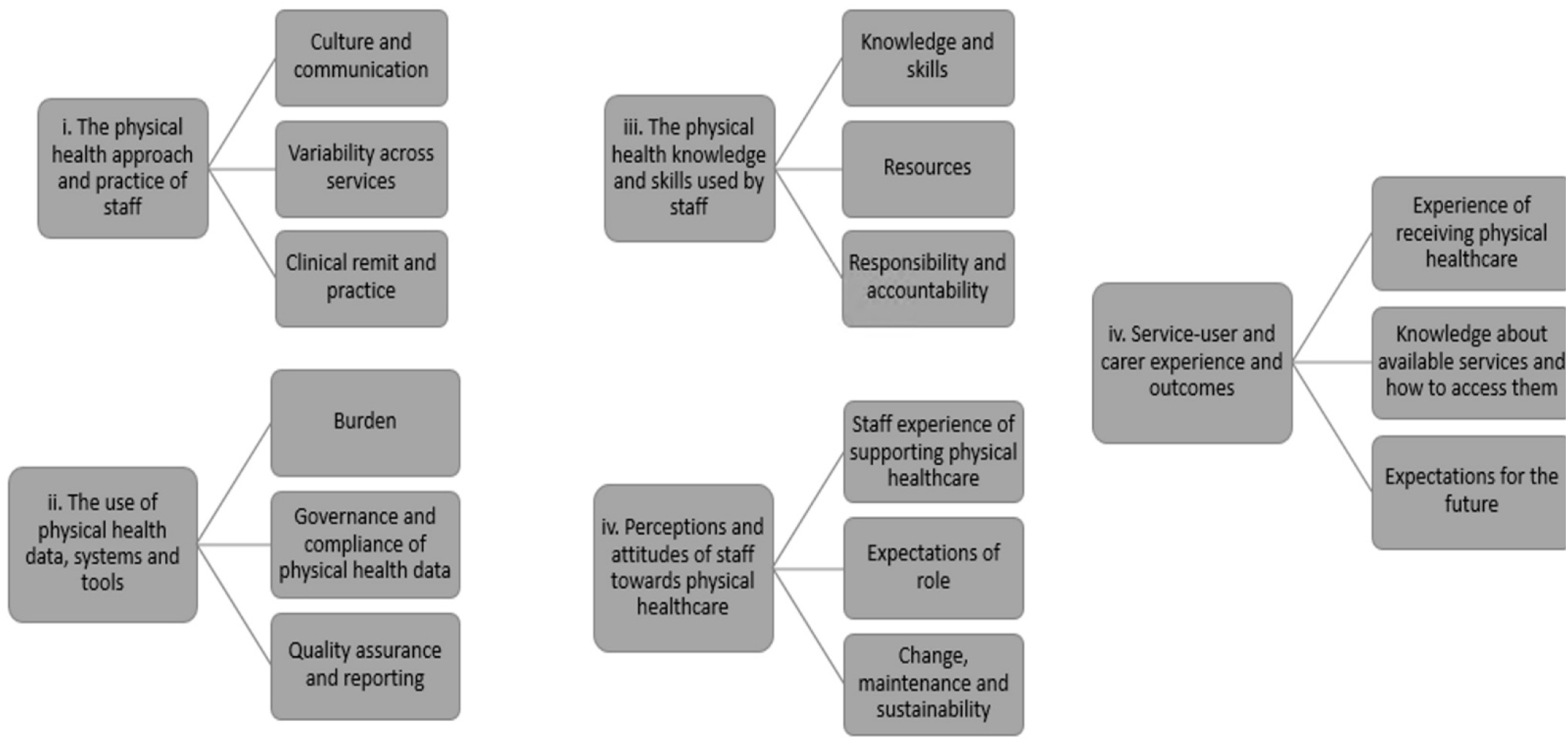
Schematic representation of the five main themes and related subthemes that were identified from the data set.

## 5 Results

### 5.1 Demographics

50 participants took part in the study. 38 were clinical staff, eight were service users and four were carers. We collected demographic data from 22 clinical staff and all service users and carers. Participant demographics are shown in Tables 1 and 2.

**Table 1.** Demographics of clinical staff.

| Demographics of clinical staff who provided their demographic data (N=22) |  |
| --- | --- |
| Characteristics | No. participants |
| *Age* |  |
| 20-29 | 2 |
| 30-39 | 2 |
| 40-49 | 7 |
| 50-59 | 9 |
| 60-69 | 2 |
| *Gender* |  |
| Male | 7 |
| Female | 15 |
| *Ethnic background* |  |
| White British | 13 |
| White Irish | 1 |
| White Other | 2 |
| Black British | 1 |
| Black African | 3 |
| Asian British | 2 |
| *No. years working within mental health services at SLaM* |  |
| 3-5 years | 7 |
| 6-10 years | 2 |
| 11-15 years | 3 |
| 16-20 years | 2 |
| 21-25 years | 5 |
| 26 years or more | 3 |
| **Missing data** | **16** |

**Table 2.** Demographics of service users and carers.

| Demographics of service users and carers (N=12) |  |
| --- | --- |
| Characteristics | No. participants |
| *Age* |  |
| 30-39 | 2 |
| 40-49 | 2 |
| 50-59 | 3 |
| 60-69 | 2 |
| 70+ | 3 |
| *Gender* |  |
| Male | 4 |
| Female | 8 |
| *Ethnic background* |  |
| White British | 8 |
| White Irish | 1 |
| Black British | 1 |
| Mixed British | 1 |
| Asian British | 1 |
| *No. years accessing mental health services at SLaM* |  |
| 2 years or less | 2 |
| 3-5 years | 5 |
| 11-15 years | 2 |
| 16-20 years | 2 |
| *Mental Health Conditions* |  |
| SMI (Psychosis, including Schizophrenia, Schizoaffective Disorder, Bipolar Disorder and Depressive Psychosis) | 11 |
| Post-Traumatic Stress Disorder (PTSD) | 1 |
| *Neurological Disorders* | 2 |
| *Physical Health Conditions* |  |
| Obesity | 2 |
| Physical health effects of medication | 8 |
| **Missing data** | 0 |

The mean age of participants was 51, with most participants being female. For the staff we had demographic data for the largest ethnic group was White British, followed by Black African, which is proportional to the workforce. Most participants had worked in the Trust for at least three years. Amongst service users and carers the largest ethnic group was also White British, followed by Black British, Mixed British, and Asian British, with one participant per non-white ethnicity. All service users, except for two participants, reported having a diagnosed SMI and had accessed CMHT services at the Trust for 12 months or more.

23 interviews were completed with clinical staff from a range of teams and professional roles. Eight focus groups were held: four with clinical staff (15 participants), two with service users (eight participants), and two with carers (four participants).

### 5.2 Findings

35 hours of interview and focus group recordings were fully transcribed and submitted for analysis. Five main themes and related subthemes were identified (see Figure 1). These themes were mapped against the five main areas of interest (themes) (see 1.1). Sections 6.2.1-6.2.4 report on clinical staff experiences and section 6.2.5. reports on the experience of service users and carers.

#### 5.2.1 Physical health approach and practice of staff

##### 5.2.1.1 Culture and communication

All participants identified with the importance of addressing physical healthcare, however, there was variability in awareness of the Trust’s vision and strategy. For example, some participants, mostly clinical managers, were aware the Trust has a vision for better whole-person healthcare, often referring to the Trust strategy Aim High, Changing Lives (2021-2026) (15) but were uncertain of how to embed this into teams and everyday practice. Participants consistently reported being unclear on policy and expectations for physical healthcare practice across the organisation. Some participants in frontline roles suggested staff involvement in developing visions and strategies to inform change initiatives would help with this lack of clarity.

> *“At this stage, I don’t know if [the policy] has been put into practice yet. It feels like an idea, and it sounds positive, but what does that look like on the ground? How is it impacting the actual delivery of care? We can have ideas around what things should look like, but how has that changed the way care is being delivered?” – P1*

Participants’ experiences of communication from the Trust regarding physical healthcare varied largely based on their role and the borough in which they worked. Participants who had experienced positive communication described clear guidance from managers and regular opportunity to feedback on their experiences. Participants who had encountered difficulties in communication were unclear about the both the organisation’s role and their individual remit, but also reported a lack of opportunities to discuss experiences of physical healthcare with colleagues or managers.

> *“We’re trying to bring in a structure for communicating things, so we’re actually communicating information that needs to be handed over in a clear and concise manner. I know I’ve sat in meetings where people talk round and round, and you don’t actually get a sense of what’s happening. I think it’s [providing] a safe space for people to say, ‘I’m not sure about this,’ and feel supported so they’re not carrying things… it’s about team ownership…working together as a team*.*” – P2*

##### 5.2.1.2 Variability across services

Participants discussed variability in how individual services and teams approached physical healthcare in routine practice. Some participants in managerial roles identified that they had the confidence to use their personal agency to develop plans locally, whereas others felt they needed more support from strategic leaders to do this. Some participants reflected on variable progress in the approaches to physical healthcare in their localities. All participants recognised the need for localised plans that respond to the needs of the local population, however many noted this could also be a barrier to consistency across the organisation. Enablers included having a more structured and coordinated implementation plan for physical healthcare to ensure consistency between teams, and regular forums for frontline staff and managers to share best practice and develop common solutions.

> *“There’s confusion across all the different boroughs, we are all supposed to be doing the same thing, [but] some boroughs have got their own physical health team, others don’t, so my confusion is what does the central team do for us? Things like that*.*” – P3*

Most participants reported variability of approach was a barrier in how physical health checks were approached across services which led to an uncoordinated approach to engaging service users and engagement with primary care. Some participants expressed confusion about what physical health monitoring should look like whereas others were confident about what was expected and were keen to develop and share best practice with others.

> *“At the moment we know all the different teams work differently. We’re trying to look through the data, see where the good practice is, see where the maybe not so good practice is, and then target those specific areas*.*” – P4*

##### 5.2.1.3 Clinical remit and practice

Participants in frontline clinical roles reported a lack of clarity on who is accountable for providing physical healthcare which was largely perceived as the responsibility of primary care. Some participants were concerned that if mental health staff complete physical health checks, aspects may be missed, or primary care may perceive those needs are being addressed in secondary care services when they cannot be. Participants in frontline roles suggested that to mitigate confusion staff in SLaM need to be clear about their roles and responsibilities when providing physical healthcare and this should be shared with colleagues in primary care, as well as service users and carers.

> *“I think sometimes the right hand doesn’t know what the left hand’s doing…the GP thinks mental health services are doing it, the mental health services think the GPs are doing it. I mean, there’s an argument…for both sides to do that*.*” - P5*

#### 5.2.2 The use of physical health systems and tools

##### 5.2.2.1. Burden

Most participants in frontline staff roles reported barriers to using multiple processes and systems for recording physical health including capacity, burden, and knowledge. The main concern was a lack of confidence in what data should be recorded, how, why, and where. Most participants felt greater clarity was needed on why these data are collected in mental health services, and how they can be used effectively in mental health care provision and planning.

> *“[Using multiple systems] has increased my personal workload because, every time I see the patients, I go through the physical health, I go through the records, and again it’s other issues – so records are not always up to date…so, it increases the workload…I need to screen to make sure the patient has the kind of physical health investigations as they should*.*” - P6*

##### 5.2.2.2. Governance and compliance of physical health data

Governance and compliance of physical health data capture, use and reporting was variable across teams. Some participants in frontline roles commented that the use of multiple systems to capture service user physical health data created difficulties in practice, such as poor integration of and continuity across care records. Some participants in managerial roles reported difficulties in navigating various data systems to report physical health data accurately and to a consistent standard. Streamlined systems or greater interoperability between systems, clearer guidance, training, and refresher training, as well as clear access to support in the event of concerns with data completion or reporting were seen as enablers.

> *“A system that can be put in place to help us prioritise and continue to monitor physical health issues I think that would be helpful as well, because currently we don’t really have a system to monitor that and keep track of things*.*” - P7*

##### 5.2.2.3. Quality assurance and reporting

Many participants in frontline roles wanted simpler reporting systems and tools within routine practice to support how they monitor and report on the physical health of their service users. Some participants in managerial roles identified concerns about how to effectively report physical health data and how to support their staff to collect accurate data that can inform good quality reporting processes. Most participants in managerial roles were concerned about quality monitoring and reporting procedures physical health, for example, many reported issues with reporting effectively upwards and using data to inform improvements in services on the ground.

> *“I think, for me, data’s helpful but, unless you’ve got the context, you can’t place it, you can’t understand it. You go to any quality and performance meetings at the board level, they want data, but the data is only telling you one side of the story. It’s not giving you the barriers or enablers as to why your results are what they are. I think we don’t look at the qualitative stuff. I know we’re looking at bigger numbers and it’s a performance thing, but I think that is a really key part*.*” - P8*

#### 5.2.3 The physical health knowledge and skills of staff

##### 5.2.3.1. Knowledge and skills

All participants were aware of and had completed the mandatory Trust training on physical health but perceived it as limited (both in terms of content covered and duration) and requiring further development if physical healthcare is to be given more prominence in routine practice. Most participants felt role-specific training would ensure specific learning needs are identified and met. Participants in managerial roles identified training on how to manage team approaches to physical healthcare practice would be helpful.

> *“Staff in the community are coming from a different background. It could be nursing staff who know how to do basic things like blood pressure, etc*., *pulse, respiration, etc. And there are social workers who don’t have a clue what we’re talking about. …I think, when we target the training, the training should be targeted at different groups in the community mental health team*.*” – P9*

##### 5.2.3.2. Resources

Some participants felt there was a lack of resources (e.g., information, time, and equipment) to support physical healthcare within Trust services. Participants felt physical healthcare could be difficult to prioritise alongside other targets, or when staffing is limited. Some expressed often being without immediate access to key equipment or estates and having to source alternative options to be able to administer physical healthcare. Some participants also sought more accessible information on common physical health problems that could be given directly to patients, such as self-help resources. Participants felt that a central resource with information that could be shared directly with service users and carers would also be helpful to assist with relevant signposting and referrals. Many participants also mentioned the value of existing knowledge from colleagues who were able to share their experiences to support others, whether in their roles, or as champions of physical health.

> *“If we had the staff, if we had the resources…but…because we haven’t got the resources…unfortunately, because of low resources and staff shortages, we haven’t got enough clinical rooms or substantial volumes of equipment and staff to facilitate more in the way of physical health [alongside routine mental health care]*.*” – P10*

##### 5.2.3.3. Responsibility and accountability

Participants in all roles were interested and motivated to learn more about how to support physical healthcare but there was uncertainty about who was responsible or accountable for providing physical healthcare in the Trust. Some participants in frontline roles recognised their lack of experience in providing physical healthcare but would support provision of it if given appropriate training or support. Enablers identified by participants included: role clarity, opportunities to continuously review knowledge and skills post-training e.g., through refresher training or skills sharing, and reminders within teams about who is available to support physical healthcare (e.g., champions). For managers there was an interest in establishing a competency framework to support staff and ensure consistent individual and team competency.

> *“At the moment we don’t formally assess on people’s competency on physical health… I think that would help, as long as we provided the training, we provided the opportunities for people to use that, then we can then assess people to say, ‘OK, how is it going?’ and then that would hopefully give us the gaps that we can try and fill in. That would be good…we do for medication competencies to make sure people are practicing safely, so I suppose that would be a good thing to have*.*” – P11*

#### 5.2.4 Perceptions and attitudes of staff regarding physical healthcare

##### 5.2.4.1. Staff experience of supporting physical healthcare

All participants were highly motivated to support the physical health of their service users. Most participants in managerial roles reported an increase in awareness about physical health problems service users may face and noted a general change in attitude amongst staff to talk more openly about their experiences, to share knowledge, and to support colleagues in day-to-day practice. Most participants in frontline roles were aware of at least one person within their team who was knowledgeable about physical health they would feel confident to approach if they had a question.

> *“I think it’s a lot higher on the agenda. I just think that there did need to be additional support to support people with it. I think that that’s quite important*.*” – P12*

##### 5.2.4.2. Expectations of role

All participants sought clarity over their role in supporting service user physical health. Many participants in frontline roles reported service users did not seek specific physical health support, and where it was discussed, it was usually regarding issues that arose as a result of a mental health problem e.g., side effects from taking psychotropic medications. As a result, some participants expressed confusion about how much they were expected to know about physical health within their roles. Most participants in frontline roles expressed uncertainty in how to discuss physical health problems with service users as they were not sure what their role was. Some participants in managerial roles also lacked confidence in how to talk about physical health with their staff and how to set clear expectations or give appropriate guidance.

> *“If you haven’t done a physical health screen in a year, you might not feel very confident… if it goes off the radar, then the confidence level changes, and then… that can contribute to why somebody might feel a little bit less confident or enthusiastic even about doing a physical health care screen*.*” – P13*

##### 5.2.4.3. Change, maintenance and sustainability

Participants expressed that change in the way physical healthcare is approached and practiced within CMHTs was needed to improve overall care outcomes. Additionally, most participants reported innovation fatigue with the number of changes to policy, procedure, and practice experienced in recent years. Most participants reported interest in developing physical healthcare interventions and pathways to support physical health but wanted a more coordinated plan from the Trust to be able to do so. Participants in managerial roles suggested a greater focus on maintenance and sustainability of initiatives would better support staff to deliver a consistent model of healthcare more confidently. This could be strengthened with a focus on improvement of existing interventions rather than constant reinvention or major organisational change which tends to be disruptive to staff learning, practice, and service user care.

> *“It would be good to see outcomes from interventions that happen and any sort of changes. I’m sure that sort of thing will be available, reports and things, rather than just numbers going up, but just what that actually means and any feedback from the Trust as a whole about what’s been happening, about any changes that we’ve had and what the differences it makes in outcomes, would be good to see. I think that always helps people to see that it’s actually meaningful in every person’s life*.*” - P14*

#### 5.2.5 Service user and carer experience of accessing physical healthcare

##### 5.2.5.1. Experience of receiving physical healthcare

Service users and carers reported that in their experience mental health staff are not confident in talking about physical health and this impacted on their confidence to disclose physical health problems, which limited support-seeking for physical health problems when under mental health services. Carers experienced issues with mental health staff not understanding the possible physical health problems that can affect people with long-term mental illnesses, and as a result most carers said they would not direct physical health support enquiries to a mental health practitioner.

> *“Well, if it’s a lot of physical health issues, obviously they’re out of scope; they don’t have experience; they don’t know. They don’t want to give advice. They don’t have enough experience and enough knowledge for certain physical health, so they can’t give that advice for it. So, what they’ll do is they’ll discuss with myself, or the doctor, and we’ll discuss with the GP to make a plan or signpost us away from secondary care services*.*” - P15*

##### 5.2.5.2. Knowledge about available services and how to access them

Service users did not perceive mental health trusts as providers of physical healthcare. They would seek advice from their GP if they had a physical health problem.

> *“…it’s about continuity - where primary care or the GP have a similar level of knowledge to what SLaM do, I think there is a huge disparity. GPs largely seem to be the physical health experts, SLaM largely seem to be the mental health experts, and never the twain shall meet, which is usually problematic for service users navigating through an already complex system*.*” – P16*

Service users did not expect to receive physical health support from their mental health team and wanted to know what support was available. Service users were concerned that the physical health data captured by their CMHT was not currently being used to inform their care, or additional support they may be entitled to access. All service users said if they were better informed, and more involved in their care, they may have a better understanding of what support is available and how they can access it.

Carers conveyed frustration that physical and mental health needs are separated and accessed via different services. Carers suggested it would be helpful if secondary care services had knowledge of how to check for signs of poor physical health amongst adults with SMI and could give guidance on how to prevent symptoms worsening. Carers said if they knew what support was available within secondary care, they too could encourage service users to ask for it or to enquire on their behalf.

##### 5.2.5.3. Expectations for the future

Overall, service users wanted more clarity about what physical healthcare was available from secondary care, how this could be accessed, and how this could support their care outcomes. Carers sought greater ownership and transparency from secondary care about what they provide, and better communication with service users about options to access, or to review it.

> *“I was told recently that I’m very anxious - that was news to me, and I couldn’t relate. I’m beginning to notice that physically more within my body and was realising the struggle to get into the day…I’m feeling strange and unsettled. I introduced that to the mental health side, and it’s like, ‘that’s trivial*.*’ I mentioned it to the GP, and it’s again trivialised, not connected with. The broader thing is, you’ve got mental health, so the physical health impact isn’t taken into account. It’s not coordinated at all, it’s kind of clunky*.*” – P17*

All service users and carers agreed integrated care was important for the future of mental health services and wanted clarity about what this means in everyday practice.

## 6 Discussion

In this service evaluation we found that there was high staff motivation to support the physical healthcare of adults with SMI, but staff wanted more clarity about the Trust’s vision and strategy, and importantly, how this aligns to their roles and should benefit service user care. Staff had clear views of what would support them including clear operational guidance, a comprehensive training programme, clarity around roles and responsibilities, clear physical health leadership, and collaboration from colleagues. Staff also wanted to feel part of decision-making about physical health and to contribute to a best practice approach across the organisation. Our findings add to existing research. For example, we identified the impact on staff of having unclear guidance, limited knowledge, and a lack of resources when administering physical healthcare, building upon the findings of Small (2017) (2) and Gray (2017) (3). We also found staff experience burden when they perceive themselves as lacking in confidence or without appropriate skills to practice physical healthcare which echoes the work of Papachristou et al (2019) (4). Yet our findings go further and show the need for clearer leadership in physical healthcare, supported by structures that facilitate a culture anchored in robust training and policy. Work by Belling (2011) (16) has shown consideration of culture and leadership when implementing change influences staff attitudes and practice, indicating this as a valuable finding from our study.

Service users did not expect mental health providers to offer physical healthcare but wanted a more integrated and coordinated approach across primary and secondary care and sought clarity on exactly what physical health support CMHTs could offer alongside their GP. Carers stressed the need for transparency with the service user about what physical health information is being collected, how this will support their care, and how they can be involved in using that information to support care planning and improvements in practice. A recent review suggested the views of service users towards interventions to screen for and treat physical healthcare problems in secondary care are largely overlooked in research and could be hindering effective implementation in practice (17). This review reported that authentically including service users in physical healthcare evaluation can support policy being implemented in a tailored way that truly meets the needs of service users. Moreover, there is a need to communicate better with service users about their physical health, what they can expect to receive from services and how this will support their journey of care.

For carers, we identified similar findings to the studies conducted by Onwumere (2018) (8), regarding the burden of poorly coordinated care being passed onto carers. However, our findings went further, as carers in our study wanted to be informed by healthcare providers about what care options are available. Carers felt having the opportunities to convey their needs was vital, and wanted clarity over available support, as well as better coordinated and equitable care for their loved ones. They felt that that the absence of this hindered the experience, care and recovery of the service user they were supporting.

More coordinated and integrated care is the aim of the Community Mental Health Transformation Programme (18). However, a recent paper by Hannigan et al (2018) (19) suggested healthcare providers need to consider what their expectations are for improved coordination and how changes in policy play out in routine practice if it is to be achieved sustainably. This review of mental health policy in practice showed effective care coordination across primary and secondary services can be achieved through strong communication, collaborative working and training. It is, however, time consuming and resource dependent which can often lead to strain on staff, resulting in outcomes such as fatigue, withdrawal, or resistance towards original change ideas or policy. This is key as poor implementation of policy not only leads to poor adoption from staff, but ultimately hinders practice that impacts service user care. Our findings reflect this thinking, as SLaM staff shared their frustrations with constant change, and wanted a focus on the maintenance and sustainability of interventions.

Overall, the broader context for staff, service users, and carers concerned the general approach of the organisation towards physical healthcare, and how this was shown in the vision, strategy, policy, culture, organisational leadership, and adoption of change initiatives within individual services. For example, staff expressed the need for greater support for leaders and managerial staff to enable them to guide and support their teams and work with partner organisations to deliver good quality physical healthcare. Equally, team leaders, managers or supervisors identified a gap in their own knowledge and practice when supporting frontline staff to realise the organisations vision for physical healthcare and to make appropriate decisions in routine practice. A study by Singh (2000) (20) highlighted the significance of having clarity of purpose, a shared vision and frequent review of team operations to achieve effective team cultures, behaviour, and outcomes. This paper shows that if secondary care services plan to support the physical health of service users, greater consideration is needed by the organisation towards the implementation of that vision, and the culture, processes, and systems required to ensure good quality physical healthcare.

### 6.1.1 Strengths and limitations

#### 6.1.1.1. Strengths

The research team’s diverse membership of clinicians, researchers, and experts by experience enabled greater collaboration over research design and delivery. The involvement of the evaluation team ensured a range of perspectives (including that of clinicians, academics, and service users) informed decision-making throughout the evaluation process - including in setting up the scope and aims of the work jointly with the Trust at the onset of the evaluation, which enhanced the utility of the findings. High engagement from staff, service users, and carer participants enabled the researchers to select from a large pool of interested participants from a range of different professional roles. This meant that there was a healthy sized sample for this qualitative study, which was both professionally diverse amongst the staff participants, and demographically diverse amongst service user and carer participants. As a result, we were able to conduct all planned interviews and focus groups and obtain rich and varied experiences based on current practices in adult CMHTs.

#### 6.1.1.2. Limitations

16 staff participants declined to complete the demographic questionnaire provided, which means that despite our efforts to pre-select a demographically diverse sample, we are unable to accurately report this. Therefore, we are unable to determine how representative our final sample is of staff working within the Trust. Further, due to the nature of the methods used, we are unable to generalise beyond the context of the services and participants who took part in this work.

### 6.2 Implications for policy

These findings show potential gaps in guidance on how physical healthcare is being approached within community services. In 2014, NICE set out recommendations for the completion of physical health checks in secondary care services. Yet the findings of this study suggest our understanding of what is required and how it should be delivered has evolved since this guidance was published. There is therefore a question of whether additional guidance is required to support mental health Trusts in their approach to physical healthcare, both in the interventions they provide but the follow-up care that is offered to service users as well. Moreover, in light of the recent transformation of CMHTs across England, this could be an opportunity to consider how guidance on physical healthcare applies and if basic checks are sufficient to support service users in the future.

### 6.3 Implications for clinical practice

To improve the experience of staff, service users and carers, the vision regarding physical health needs to be better communicated and understood. There needs to be better training for staff, more clearly defined roles and responsibilities, and greater opportunities for staff, service users and carer involvement in defining how physical health is supported. The research team aims to support the Trust with this using Implementation Science methodologies that help with change management. These methodologies could also be helpful for other Trusts.

### 6.4 Implications for research

The findings could be used to explore whether similar themes are true and generalisable to other Trusts that offer mental health services. The research team aims to use the findings from this evaluation to develop a framework that could support the Trust to consider physical healthcare practice in community settings. Further research could be completed to evaluate the use and impact of this framework in other Trusts to support a more consistent approach within CMHTs generally. Importantly any future research must prioritise the involvement of staff, service users and carers to ensure their experiences are embedded into and inform future initiatives.

### 6.5 Conclusion

Mental health staff recognise and are motivated to provide support for the physical health of adults living with SMI. The findings presented in this paper provide insight into common barriers and facilitators faced by staff, service users, and carers when providing or accessing physical healthcare within adult CMHTs. This evaluation study has led to better understanding of the impact on care outcomes of service users and will help to develop more effective ways to address and improve them. Moreover, we have explored the role of mental health staff and what may need to change for them in how they work within teams, and how they interact with other parts of the organisation to improve physical healthcare for people with SMI. The findings indicate that what is needed is a more comprehensive and sustainable approach to physical healthcare provision in CMHTs.

## Data Availability

Anonymised transcripts of interviews and focus groups are available from the evaluation group upon request.

## 7. Resource Identification Initiative

Not applicable

## 8. Life Science Identifiers

Not applicable

## 9. Conflicts of interest

NS is the director of the London Safety and Training Solutions Ltd, which offers training in patient safety, implementation solutions and human factors to healthcare organisations and the pharmaceutical industry. The other authors have no conflicts of interest to declare.

## 10. Author contributions

GT, JW, RM, FG, NS designed the service evaluation. GT undertook all data collection with support from JW, RM, JT and LO. Analysis was undertaken by GW, JW, RM with support from ES, NS, FG, NSt, JT and LO. The manuscript was written by GT and JW with comments from RM, NS, FG, ES, SC, NSt, JT and LO.

## 11. Funding

The evaluation was funded by the Maudsley Charity. NS and FG’s research is supported by the National Institute for Health Research (NIHR) Applied Research Collaboration (ARC) South London at King’s College Hospital NHS Foundation Trust. NS is a member of King’s Improvement Science, which offers co-funding to the NIHR ARC South London and is funded by King’s Health Partners (Guy’s and St Thomas’ NHS Foundation Trust, King’s College Hospital NHS Foundation Trust, King’s College London and South London and Maudsley NHS Foundation Trust), and Guy’s and St Thomas’ Foundation. ES is supported by NIHR ARC Wessex. RM is an ICA Pre-doctoral Clinical and Practitioner Academic Fellow supported by Health Education England and the NIHR Research. The views expressed in this publication are those of the authors and not necessarily those of the NHS, the NIHR, the ESRC or the Department of Health and Social Care.

## 12. Acknowledgements

The authors would like to thank all participants who took part in the evaluation, as well as the experts by experience and clinical advisors who supported its completion. Additional thanks to our Project Sponsor, South London and Maudsley NHS Foundation Trust, and particularly to Deputy Chief Nurse, Helen Kelsall, and colleagues via the SLaM Physical Health Quality Centre who supported delivery of this evaluation. Finally, the authors would like to thank their Project Funder, the Maudsley Charity, for their continued support to complete this evaluation on behalf of the Trust.

## 13. Supplementary Material

Copies of the interview and focus group topic guides used for clinical staff, service users and carers has been included in the supplementary material.

## 18. Supplementary Materials

### 18.1. Clinical Staff Interview and Focus Group Topic Guide

#### 1. Physical health approach and practice

##### 1.1. Identify the standard journey for a patient accessing the community mental health team

###### Prompts

- *How are the majority of patients referred to your team? (e*.*g*., *primary care, social care)*.
- *What is the standard approach for patients that are referred: screening, assessment, allocation? Might physical health needs be identified at any of those stages? If so, what would typically happen?*
- *Would you accept a referral for a patient where physical health needs were identified as contributing to their mental health? If no, why not? If yes, how would you explore the comorbid concerns as a team?*
- *Does physical health feature in the decision when you allocate a patient to a Care Coordinator? How?*

##### 1.2. Identify which roles are likely to support patients with physical health problems

###### Prompts

- *Do you have any nominated champions of physical health that support you and the team? If yes, how are they appointed and what does their role involve? If no, why not?*
- *Do you work with teams in other organisations, such as primary care, social care, other hospitals, housing, around physical healthcare? How? Are there any challenges around this? What do you find helpful?*
- *Where do you go when you encounter a problem with patient’s physical health?*

##### 1.3. Identify physical health priorities for the service

###### Prompts

- *How much do you think the team prioritises physical health? Why?*
- *How are priorities set in the team? (E*.*g*., *are they discussed or mandated?) Are you asked to meet any targets/reporting around physical health? How are these communicated to you?*
- *How often does physical health come up in conversation with colleagues? Where does this come up? Prompt: team meetings, supervision*

##### 1.4. Explore perceptions of physical healthcare practice within the team and across the Trust

###### Prompts

- *Does your locality have its own strategy for physical health? What works well / not so well about your local approaches to physical health? Does this align to the Trusts?*
- *Do you know what the Trusts position on physical health is? Do you know where to find it?*
- *The new strategy states that physical health and mental health care will be treated equally. Do you agree with this approach? How confident are you that the Trust can realise this vision, and that these priorities will actually make a difference to patient care?*
- *Based on your experience do you face any particular challenges when putting into practice the physical health priorities of the Trust? What barriers do you foresee for physical healthcare in the future?*

#### 2. Use of physical health systems and tools

##### 2.1. Confirm the main databases, systems and tools used by the team (use checklist as prompt)

###### Prompts

- *What is the main patient record system used by your team? How do you use this system when identifying/recording physical health needs?*
- *Does the system enable you to provide better physical healthcare? Is there anything you feel could improve experience for you/ your staff?*
- *Do your staff use any specific tools to help patients with physical healthcare needs?*
- *What experiences have you had / found your staff have had when using these tools?*
- *Do you use any additional systems/tools to identify, record, or report about physical health? If so, what do you use, and what is the staff experiences of using these?*

##### 2.2. Communication of patient needs

###### Prompts

- *Are there systems in place to be able to communicate easily with other professionals? If yes, which professionals do you work with (e*.*g*., *inside SLaM, primary care, acute care). Are there any barriers/facilitators to this?*
- *Is there anything else you are doing in your role or as a team/service to champion physical healthcare that you think other teams could learn from?*

#### 3. Physical health knowledge, skills, and training

##### 3.1 Identify the main problems experienced

###### Prompts

- *What types of physical health problems do you work with your patients to manage?*
- *Do you feel that you have the adequate knowledge, skills, and training to deliver what is being asked of you by the patient/carer/team/Trust?*
- *What impact has this had on the way you approach or provide healthcare to your patients?*
- *Has it changed any interventions or how long you support a patient?*
- *Do you think staff feel able to engage confidently with patients about their physical health? Are there any barriers to this? What would help improve it?*

##### 3.2. Explore barriers and facilitators to knowledge and skills

###### Prompts

- *[Anything in addition to what we have already covered] What do you think you would need to be able to respond to physical health concerns? Prompt: skills, time*
- *What gaps are there? What else would you like staff to know? Why?*
- *What efforts have been made within your own team to address gaps or issues with knowledge and skills e*.*g*., *physical health leads, physical health forums etc. How helpful have these been, and why?*

##### 3.3. Explore training experience

###### Prompts

- *What training do you ask your staff / are you asked to do when joining the team? Prompt: part of induction, reviewed at appraisal, Mandatory Level 1 training*
- *Are there any additional tools (e*.*g*., *leaflets, apps etc*.*) that could help staff?*
- *Do you feel your team does anything specific to develop staff knowledge and skills around physical health? E*.*g*., *clinical / case reviews, business meetings, management rounds, supervision etc*.
- *[If not mentioned previously]. Is physical health competency or activity reviewed as part of your supervision? To what extent do you find this helps you to manage your development in this area?*

#### 4. Physical health attitudes, perceptions, and experiences of staff

##### 4.1. Identify the personal experiences and perspectives of the interviewee towards physical health

- *How important do you feel physical healthcare is in your role? Why is that? Any personal experiences you could draw upon?*
- *What has been your personal experience of supporting SMI patients with their physical health? What obstacles have you faced?*
- *What do you think the role of mental health staff should be in supporting people’s physical health? How successfully do you think staff achieve this in their roles now?*
- *Is there anything you hope to/would like to/think could be improved or changed about how physical healthcare is approached within your team / at SLaM? What do you think would be important when implementing these things?*

#### 5. Other (value added)

##### Prompts

- *What would you like to see from the Trust when looking at physical healthcare for the future?*
- *What more could the Trust do to engage staff around physical healthcare in the future?*
- *Examples of good practice*
- *Lessons that can be learned/shared (case study template)*

#### 6. Anything other business

##### Prompts

- *Areas for future research on physical health*
- *Anyone else that would be useful to speak to*.
- *Anything else that the interviewee feels has been missed and anything that they did not get a chance to discuss fully*.

##### 18.2. Service-User and Carer Focus Group Topic Guide

**Topic 1:** Exploring your experience of physical healthcare when you have accessed an adult community mental health team.

- **Broad question:** What has your general experience of receiving physical healthcare been like when accessing SLaM Adult Community Mental Health Teams?
- **Narrow question:** Did you experience any problems when receiving physical healthcare? What worked well? What could help improve it?

**Topic 2:** Your perception of the knowledge and skills that mental health staff possess regarding physical health, and areas where you feel this could be improved.

- **Broad question:** To what extent do you think Adult Community Mental Health Team staff are adequately knowledgeable or trained on physical health issues?
- **Narrow question:** What knowledge, skills or training do you think staff could further benefit from to support adults with mental illness who have additional physical health problems?

**Topic 3:** The systems and tools that mental health staff use within the community mental health team to identify, monitor, record or communicate about a patient’s physical healthcare needs (e.g., physical health questionnaires, physical health referrals).

- **Broad question:** Do you know why your CMHT holds data on your physical health needs? What difference do you think this makes to your care?
- **Narrow question:** How do you feel we engage with you about your physical health, using the information that you provide? How do you think the way we hold and share your data around physical health could be improved?

**Topic 4:** The team, service, and organisational approach towards physical healthcare and in particular, the culture this creates amongst staff, patients and carers, and the extent to which this aligns to the Trust’s strategic vision for physical healthcare.

- **Broad question:** As a service-user, to what extent do you understand the physical health priorities set out by the Trust? How much does this influence your engagement with SLaM for mental and physical health support?
- **Narrow question:** What do you think the future of physical healthcare at SLaM should look like or involve?

## Notes

### Author Declarations

Approval was obtained from Clinical and Information Governance professional leads at the South London and Maudsley NHS Foundation Trust (on 14th March 2022). All participants involved in the study were briefed prior to taking part and given written information about how their data would be used within the evaluation process and as part of any final publications. Participants provided written informed consent to participate in the study. Participation was voluntary and withdrawal was possible at any stage.

